# Protocol for systematic review of measurement of fatigue in people with cerebral palsy

**DOI:** 10.1101/2021.07.20.21260898

**Authors:** I. M. Dutia, R. Eres, S. M. Sawyer, L. Johnston, D. Reddihough, S. Cleary, D. Coghill

## Abstract

**Background:** Fatigue is a common problem for people with cerebral palsy (CP), which adversely affects health-related outcomes. The emerging body of literature on fatigue in people with CP is characterised by substantial heterogeneity in assessment methods. To date, a systematic analysis of the measurement methods, and appraisal of the tools used to measure fatigue in people with CP has not been carried out. The aim of this review is to use The International Classification of Functioning, Disability and Health (ICF) as a framework to categorise and appraise current methods for measuring fatigue in people with CP.

**Methods and Design:** Literature searches will be conducted in MEDLINE, PsycInfo, CINAHL, Web of Science and Cochrane databases. Studies will be included in this systematic review if they purport to measure any type of fatigue in people with CP of any age through original research, are written in English and are published in peer-reviewed literature since 1980. From included studies we will extract the assessment methods used to measure fatigue, and demographic and clinical characteristics of the sample of people with CP. We will then provide a narrative synthesis of the type and ICF domain of fatigue purportedly measured and critical appraisal of the assessment methods used.

**Discussion:** This review will summarise current methods for measuring fatigue in people with CP. Critical appraisal and systematic categorisation of assessment methods will allow us to identify areas for further research on the domains in which fatigue occurs, and the role of fatigue in relation to clustering with other symptoms. Findings are expected to guide future assessment of fatigue in people with CP.

## Background

Cerebral palsy (CP) is an umbrella term for a heterogeneous group of neurological disorders, caused by a non-progressive injury to the developing infant or foetal brain ^[1]^. Impaired motor control is the defining feature of CP and is associated with secondary musculoskeletal problems including changes in muscle tone, reduced strength and range of movement ^[2]^. Physical impairments may present in combination with disturbances in cognition, vision and communication, which can all negatively impact independent participation in society ^[3]^.

Fatigue is a common problem for people with CP - up to 40% of adults with CP are reported to experience higher levels of fatigue than the general population ^[4] [5]^. Fatigue is known to adversely affect health-related quality of life, independence in daily activities and functional mobility, and is experienced most frequently by those with more severe motor impairment (Gross Motor Function Classification System level II-V) ^[6] [7]^. The mechanisms underpinning fatigue in people with CP remain relatively poorly understood. This limits the development and testing of fatigue prevention strategies, which is reinforced by the absence of clinical practice guidelines for managing fatigue in people with CP.

The emerging body of literature on fatigue in people with CP is characterised by substantial heterogeneity in study design and assessment methods. This heterogeneity may be attributed, at least in part, to a lack of consensus on the definition of fatigue, which is a very broad construct. Fatigue is most commonly described in the literature within a physical domain – for example, feelings of bodily tiredness, a lack of energy for physical tasks and local muscle fatigue ^[8]^. More infrequently studies report on cognitive or mental fatigue, which describes excessive cognitive tiredness or exhaustion in response to a demanding task or significant sensory stimulation ^[9]^. Such cognitive fatigue is associated with a disproportionally long recovery time and is an atypical or pathological response to a challenging task ^[9]^. Physical and cognitive fatigue can be experienced in isolation or together. They can also occur in the context of other symptoms, such as depressed mood and pain ^[6]^. In addition to presenting with different levels of severity fatigue also has a temporal dimension. It can be acute, for example during or after physical activity, or chronic, whereby excessive fatigue is experienced over prolonged periods of time with or without an attributable cause. A subjectively experienced phenomenon, fatigue can also have objectively measurable correlates such as limitations in physical strength or cognitive capacity ^[10]^.

To advance understanding of fatigue in people with CP, it is necessary to first examine the domains in which it is reported to occur, and to critically appraise the approaches to measurement which have been utilised in studies. The International Classification of Functioning, Disability and Health (ICF) ^[11]^, provides a useful framework and standard language for the description of health-related outcomes through which we can categorise these domains and measurement methods. This type of analysis – systematic categorisation of fatigue domains and appraisal of measurement methods – will indicate how fatigue has most commonly been defined and interpreted in the literature and in which domains it has purportedly been assessed.

This protocol describes our procedures for conducting a systematic review of approaches used to measure fatigue in people with CP. Specifically, we will conduct a systematic search of the literature to identify measurement tools that purportedly measure fatigue. We will report: i) the type and domain of fatigue purportedly measured; ii) the assessment methods used; iii) the domain of the ICF in which fatigue was measured; iv) the populations of people with CP in which the measure has been used; v) the population for whom the measure was designed; vi) the extent to which the psychometric properties of the tools have been evaluated and vii) clinical utility. The findings of the review will advance our understanding of the construct of fatigue in people with CP as it is reported in the literature today.

## Methods and Design

This protocol describes methods for a systematic review of studies purporting to measure fatigue in people of any age with CP. The primary outcome will be the assessment methods used to measure fatigue, and the study will aim to critically appraise and categorise assessment methods by the type and domain of fatigue they purport to measure and by domain of the ICF ^[11]^. The protocol will be registered on PROSPERO and published on an online pre-print service. We will follow PRISMA guidelines for conducting a systematic review.

In the sections that follow we describe I) study inclusion and exclusion criteria, II) literature search protocols, III) study screening and selection processes, IV) data extraction, and V) analysis of findings.

### I. Inclusion and exclusion criteria

Studies will be included in the systematic review if they meet the following criteria: i) purport to measure any type of fatigue in people with CP of any age through original research, ii) is written in English, iii) the assessment tool is written in English and iv) was published in peer reviewed literature between 1980 and 2021.

Studies will be excluded from the systematic review if: i) they include participants with CP but where data for those participants are indistinguishable from data for participants with other disorders, ii) the publication date is prior to 1980, iii) the study is published in a language other than English, iv) the assessment tool is published in a language another than English, or v) the study is unpublished or identified in grey literature.

### II. Literature search protocols

We will develop and implement a literature search strategy for selected bibliographic databases to identify relevant literature, as described below:

#### II.I Bibliographic database search

Literature searches will be conducted in MEDLINE, PsycInfo, CINAHL, Web of Science and Cochrane databases. Search terms will be developed with the support of a research librarian and will be tailored to each database. Initial strategies will include terms that relate to cerebral palsy and fatigue. We will also use truncations (e.g., * in fatigue to obtain fatigue, fatiguing, fatigued), wildcards (e.g., ? to accommodate for difference in spelling between British and American spelling), and proximity operators to ensure the search strategies are comprehensive and robust. We will use search filters to ensure that articles retrieved meet the inclusion and exclusion criteria related to date and type of publication, and English language.

#### II.II Secondary literature search

We will also use a secondary literature search strategy to identify relevant articles from published studies that may not appear in the databases, including: i) reference lists of all included articles and ii) the title and authors of any included assessment tool.

### III. Study screening and selection process

Following the search strategy, the identified literature will be screened according to the following steps: i) search results will be imported into COVIDENCE software where duplicates will be removed, ii) title and abstract review will be carried out by 2 authors (ID and RE) and conflicts resolved via discussion, or via 3rd author (DC) if required, iii) full text review will be carried out by 2 authors (ID and RE) and conflicts resolved via discussion, or via 3rd author (DC) if required, iv) secondary searches of reference lists of all included papers, and for identified literature repeat steps i)-iii).

### IV. Data extraction

A data extraction template will be developed and used to extract data from studies included in the review. Extracted information will comprise the number of participants included in study, demographic and clinical characteristics of the sample of people with CP (including Gross Motor Function Classification System level, Manual Ability Classification System level, Communication Function Classification System level, and Eating and Drinking Classification System level where possible), and the assessment method used to measure fatigue. Two authors will pilot the data extraction template on 5-10 included studies, any discrepancies will be identified and discussed, and the data extraction template will be refined if required. Two authors will then proceed to extract information on all included studies. Any further discrepancies will be discussed and resolved via discussion or with a third author if required. Any missing data will be requested from study authors through email correspondence.

### V. Analysis of findings

We will provide a narrative synthesis of the extracted data. Findings will be presented in a way that is coherent and consistent with the extracted data. We will report: i) the type and domain of fatigue purportedly measured; ii) the assessment methods used; iii) the domain of the ICF in which fatigue was measured; iv) the populations of people with CP in which the measure has been used; v) the population for whom the measure was designed; vi) the extent to which the psychometric properties of the tools have been evaluated and vii) clinical utility.

## Discussion

This review will identify and summarise current methods for measuring fatigue in people with CP across the lifespan. Critical appraisal and systematic categorisation of assessment methods will allow us to identify areas for further research on the domains in which fatigue occurs, and the role of fatigue in relation to clustering with other symptoms. Findings are expected to guide practice, and outcomes of this review will have implications for neurological rehabilitation.

## Data Availability

All data referred to in the manuscript is available upon request.

## Notes

### Competing Interest Statement

The authors have declared no competing interest.

### Funding Statement

This work is supported by the National Health and Medical Research Council (NHMRC).

### Author Declarations

As this is a protocol for a systematic review to be conducted in the future, ethical approval is not required.

